# Rest-Activity Rhythm Phenotypes in Adults with Epilepsy and Intellectual Disability

**DOI:** 10.1101/2024.09.09.24313145

**Authors:** Nandani Adhyapak, Grace E Cardenas, Mark A Abboud, Vaishnav Krishnan

## Abstract

**Objective:** Sleep and rest-activity rhythms (RARs) are perturbed in many forms of neuropsychiatric illness. In this study, we applied wrist actigraphy to describe the extent of RAR perturbations in adults with epilepsy and intellectual disability (“E+ID”), using a cross-sectional case-control design. We examined whether RAR phenotypes correlated with epilepsy severity, deficits in adaptive function and/or comorbid psychopathology.

**Methods:** Primary caregivers of E+ID adults provided informed consent during routine ambulatory clinic visits and were asked to complete standardized surveys of overall epilepsy severity (GASE, Global Assessment of Severity of Epilepsy), adaptive function (ABAS-3, Adaptive Behavior Assessment System-3) and psychopathology (ABCL, Adult Behavior Checklist). Caregivers were also asked to ensure that subjects wore an Actiwatch-2 device continuously on their nondominant wrist for at least ten days. From recorded actograms, we calculated RAR amplitude, acrophase, robustness, intradaily variability (IV), interdaily stability (IS) and estimates of sleep quantity and timing. We compared these RAR metrics against those from (i) a previously published cohort of adults with epilepsy without ID (E– ID), and (ii) a cohort of age- and sex-matched intellectually able subjects measured within the Study of Latinos (SOL) Ancillary actigraphy study (SOL). Within E+ID subjects, we applied *k-*means analysis to divide subjects into three actigraphically distinct clusters.

**Results:** 46 E+ID subjects (median age 26 [20-68], 47% female) provided a median recording duration of 11 days [range 6-27]. Surveys reflected low to *extremely* low levels of adaptive function (ABAS3 General Adaptive Composite score: median 50 [49-75]), and low/subclinical levels of psychopathology (ABCL total score: median 54.5 [25-67]). Compared with E-ID (n=57) and SOL (n=156) cohorts, E+ID subjects displayed significantly lower RAR amplitude, robustness and IS, with significantly higher IV and total daily sleep. K-means clustering of E+ID subjects recognized an intermediate cluster “B”, with RAR values indistinguishable to E-ID. Cluster “A” subjects displayed pronounced hypoactivity and hypersomnia with high rates of rhythm fragmentation, while cluster “C” subjects featured hyper-robust and high amplitude RARs. All three clusters were similar in age, body mass index, antiseizure medication (ASM) polytherapy, ABAS3 and ABCL scores. We qualitatively describe RAR examples from all three clusters.

**Interpretation:** We show that adults with epilepsy and intellectual disability display a wide spectrum of RAR phenotypes that do not neatly correlate with measures of adaptive function or epilepsy severity. Prospective studies are necessary to determine whether continuous actigraphic monitoring can sensitively capture changes in chronobiological health that may arise with disease progression, iatrogenesis (e.g., ASM toxicity) or acute health deteriorations (e.g., seizure exacerbation, pneumonia). Similar long-term data is necessary to recognize whether behavioral interventions targeted to ‘normalize’ RARs may promote improvements in adaptive function and therapy engagement.

## Introduction

Intellectual disability (ID) can be defined as neurocognitive limitations that limit learning and independent functioning (www.cdc.gov). ID affects 1 in 4 individuals with epilepsy, where it is multifactorial in etiology. In most instances, the underlying epileptogenic structural or genetic lesion is thought to be a principal determinant, since these etiologies are frequently capable of independently derailing normal intellectual development prior to seizure onset^1^. Early life seizures (with neonatal, infantile or childhood onset) may themselves disrupt neuronal maturational processes during critical windows^2^, contributing to developmental delays that often persist despite seizure remission. Frequent focal or generalized epileptiform discharges, particularly those that occur during wakefulness, may result in a potentially remediable form of mental slowing^3,4^. A higher cumulative burden of antiseizure medication (ASM) use is also associated with lower intelligence quotients (IQ)^5^, although ASM polytherapy per se may be a surrogate marker of a more virulent and widespread epileptic network.

When epilepsy is accompanied by ID (“E+ID”), there are additional challenges to seizure management that are frequently unmet in typical adult epilepsy practices^6^. Limitations in language and cognitive function necessitate a close collaboration with either aging parents and/or rotating group home staff to achieve reasonably accurate seizure counts^6^. This population also features significantly higher rates of psychiatric and behavioral side effects to ASMs^7^, which must be prescribed cautiously, on the background of an already large set of brain-acting medications (antidepressants, antipsychotics, muscle relaxants, etc.). Swallowing difficulties and gastrostomies further limit treatment options by necessitating crushable or liquid ASM formulations. ID and epilepsy frequently presents as a triad with autism^8^, where high rates of challenging behaviors^9,10^ (“meltdowns”, tantrums, self-mutilation) often preclude any form of long-term electroencephalographic (EEG) surveillance. Counseling caregivers about newer medical or surgical therapies is imperfect, since subjects with ID are frequently excluded in initial clinical trials of efficacy^11^. And finally, the moderate to severe limitations in adaptive functioning that prohibit safe independent living lead to either institutionalization and/or the need for long-term family (or other) caregivers, who are themselves a population that is vulnerable to impairments in health-related quality of life and emotional wellbeing (by virtue of their caregiving demands)^12^. Since most cases of E+ID occur because of an early life insult (e.g., perinatal hypoxia) or inherited genetic mutation, patients and their caregivers frequently struggle with lifelong neurological disabilities. Sadly, E+ID patients suffer from high rates of premature and (often avoidable) death, with SUDEP (sudden unexpected death in epilepsy), pneumonia, drowning and status epilepticus identified as leading causes^13,14^.

As we innovate and validate the many urgently needed precision therapies on the horizon, there remains an equally urgent need for biomarkers that can objectively capture clinically meaningful improvements in both seizure and non-seizure symptoms. Compared with traditional survey-based outcome measures^15-17^, physiological biomarkers collected non-obtrusively and longitudinally provide more quantitative data, while also being agnostic to culture, language or language function. Within epilepsy, research into the utility of such wearable data streams has extensively focused on the problem of seizure detection or prediction^18-21^, with little emphasis placed on non-seizure comorbidities. In this study, we continuously measured accelerometry at the wrist (“wrist actigraphy”) in a sample of adults with E+ID to quantitatively assess abnormalities in rest-activity rhythms (RARs). Using activity “counts”^22^ (collected in 30- or 60-second epochs), wrist actograms can provide insights into RAR morphology (height, timing and robustness) through parametric techniques (e.g., cosinor modeling)^23^. Several non-parametric endpoints have also been widely utilized, including estimates of rhythm fragmentation (intradaily variability [IV]), stability (interdaily stability [IS])^24,25^ and sleep^24,26^. From large-scale actigraphic assessments of hundreds to thousands of community-derived subjects, we know that RARs display stereotyped variations by age, sex and race/ethnicity^26-28^. Similar case-control analyses have clarified the extent of RAR dysmorphology in major depression^25,29-31^, frailty ^32^ and Parkinson’s disease^33^. Within the epilepsies, two published case-control comparisons showed that adults with epilepsy display RARs with deficits in amplitude, robustness (low IS, high IV) and increased total daily sleep^26,34^, but excluded subjects with ID. Here, we similarly characterize RAR parameters from a cohort of E+ID subjects followed at tertiary care epilepsy referral center. We compare these metrics to those obtained from a previously published cohort of patients with epilepsy *without* ID (E-ID) from the same referral center^26^, and a sex- and age-matched cohort of historical control subjects^35^.

## Methods

Subjects and Protocols: This cross-sectional study is reported according to STROBE guidelines. All study methods were approved by the Baylor College of Medicine Institutional Review Board. Adult subjects with epilepsy and intellectual disability (E+ID) were recruited during routine ambulatory clinic visits at the Baylor Comprehensive Epilepsy Center between September 2022 and November 2023. We included subjects aged 18-75 with English-speaking caregivers, from whom consent to participate was obtained during routine clinic visits. We aimed to capture data from approximately 50 subjects, which is comparable to previous cross-sectional actigraphic characterizations of persons with epilepsy^26,34^. Caregivers were instructed to ensure that subjects wore the widely utilized^36-40^ Actiwatch-2 actigraphy device (Phillips Respironics) on the non-dominant wrist (or preferred wrist if hand dominance was unclear) for at least ten days. Caregivers were instructed to complete the following forms on paper-and-pencil, including (i) the single-question GASE survey^41^ (Global Assessment of Severity of Epilepsy), (ii) the Adaptive Behavior Assessment System Third edition (ABAS-3 ages 16-89) surveying behaviors critical to independent functioning, including social, daily living and communication skills^42-48^, and (iii) the Adult Behavior Checklist (ABCL), surveying psychopathology^48-50^. The ABAS-3 assesses three major domains of adaptive function (*conceptual, social* and *practical*) through 10 skill areas (communication, community use, functional academics, health and safety, home or school living, leisure, self-care, self-direction and social). Since most our subjects were unemployed, we did not include the last skill area (work). Each section comprises of 20-26 multiple choice questions pertaining to functional abilities (e.g., “Buttons own clothing”, or “states his or her telephone number), scored from 0-3 (0: is not able, 1: never/almost never, 2: sometimes, 3: always). Raw scores in each skill area were transformed to age-corrected scaled scores to obtain three adaptive domain scores (conceptual, social, practical) and a total general adaptive composite score (GAC). In contrast, the ABCL presents a total of 126 prompts as problem items (e.g., “Damages or destroys own things”), which respondents score from 0-2 (0: not true, 1: somewhat/sometimes, 2: very true/often true). Raw scores on each of 126 ABCL problem items were summed and transformed to achieve age- and sex-scaled T-scores for each of three problem areas: *internalizing* (encompassing anxious, depressed and withdrawn behaviors with somatic complaints), *externalizing* (aggressive, rule-breaking and intrusive behaviors) and *total* (incorporating, a third category of thought and attention problems).

### Control Subjects

Age, sex, body mass index (BMI) and RAR parameters in E+ID subjects were compared against two control populations. The first, “E-ID” is a published^26^ cohort of 59 adults with epilepsy recruited from the *same* clinic who were instructed to wear the *same* watch (Actiwatch-2) for the *same* duration of time (10 days). This E-ID cohort excluded pregnant patients, those with cognitive impairment sufficiently severe to preclude self-consent or self-report and those with obvious mobility limitations (hemiparesis, quadriparesis). ∼37% of this cohort were employed either part- or full-time. Individuals in the second control group (“SOL”) were selected from a publicly available annotated actigraphy data from the Hispanic Community Health Study/Study of Latinos (SOL). SOL enrolled an initial cohort of 16415 non-institutionalized subjects from Miami, Bronx, San Diego and Chicago who self-identified as Hispanic/Latino. Exclusion criteria included pregnancy, a diagnosis of narcolepsy/sleep apnea and previous/current positive airway pressure treatment^35,51^. Following an initial comprehensive baseline clinical assessment (2008-2011), 2245 subjects completed an ancillary actigraphy study, and were asked to wear an Actiwatch Spectrum Device (Phillips Respironics) on their non-dominant wrist for *7* days. The *self-reported* prevalence of common medical comorbidities (e.g., diabetes, hypertension) was recorded, but an epilepsy diagnosis was not formally ascertained. Actigraphy data and demographic variables were downloaded from the National Sleep Research Resource (www.sleepdata.org^35^). The “SOL” control group depicted in Figure 1 was made up of three or four age- and sex-matched SOL subjects for each E+ID individual. Such selected control subjects were either residing in the Bronx (∼27%), Chicago (∼25%), San Diego (18%) or Miami (30%). ∼25% did not have a high school diploma (HSD), ∼35% did have at least an HSD and the remaining 40% had greater than an HSD equivalent.

**Figure 1.**
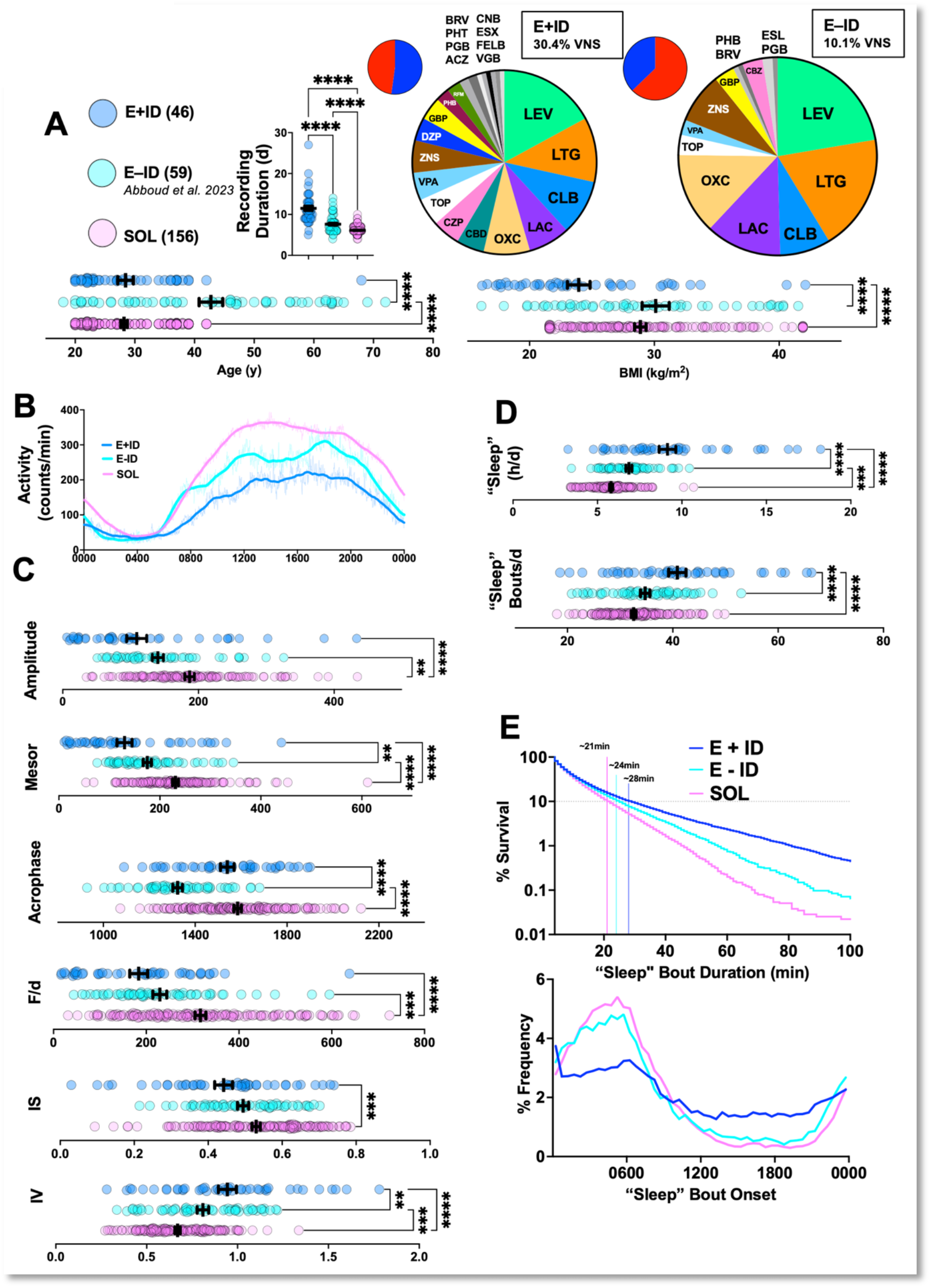
Comparing RARs in E+ID, E-ID and control subjects (SOL). A: E+ID subjects provided actigraphy recordings that were significantly longer in duration (F2,258 = 137, p<0.0001). E+ID subjects were younger than E-ID subjects (F2,258 = 51.2, p<0.0001) and had lower BMIs (F2,258 = 13.2, p<0.0001). % of subjects with active VNS therapy was higher in E+ID subjects. Pie charts of sex (blue-male, red-female) and ASM prescriptions in both epilepsy cohorts are shown. B: Averaged (thin lines), and averaged *and* smoothed (thick lines) actograms from all three cohorts. C: E+ID subjects displayed significantly lower amplitude (F2,258 = 17.4, p<0.0001), mesor (F2,258 = 34, p<0.0001), duration-normalized F (F2,258 = 20.3, p<0.0001), IS (F2,258 = 7.35, p<0.001), IV (F2,258 = 27.3, p<0.0001). D: E+ID subjects displayed the highest total daily “sleep” (F2,258 = 60.8, p<0.0001) and “sleep” bouts/day (F2,258 = 23.4, p<0.0001). E: TOP-The distribution of “sleep” bout durations shown as a survival plot using a logarithmic y-axis. Bottom: Frequency distribution histogram for “sleep” bout onset demonstrating a more limited consolidation of sleep to a main sleep period in E+ID subjects. Demographic, epilepsy-related variables and survey scores for all 46 patients. Abbreviations: LTG (lamotrigine), OXC (oxcarbazepine), ZNS (zonisamide), LEV (levetiracetam), LAC (lacosamide), CZP (clorazepate), DZP (diazepam), VPA (valproic acid), CBD (cannabidiol), GBP (gabapentin), ESX (ethosuximide), CLB (clobazam), RFM (rufinamide), PHB (phenobarbital), TOP (topiramate), FELB (felbamate), PHT (phenytoin), BRV (brivaracetam), CNB (cenobamate), VGB (vigabatrin). F/d (duration normalized pseudo-F statistic), IS (interdaily stability), IV (intradaily variability). **, *** and **** correspond to p<0.01, 0.001 and 0.001 for post hoc comparisons. Mean + standard error of the mean shown for all.

### Data Analysis and Statistics

For E+ID subjects, age, height/weight and prescription data were extracted from the electronic medical record entry corresponding to the clinic visit where consent was obtained. Ages are presented in 5-yearlong ranges (e.g., 20-24 years old) to preserve patient anonymity. Benzodiazepines were tallied as ASMs if they were administered on a scheduled basis regardless of indication (e.g., diazepam for spasticity). Age of seizure onset, seizure type, presumed etiology and documented EEG abnormalities were also obtained from chart review. For simplicity, seizure frequency was categorized as either seizure-free, sporadic, monthly, weekly or daily. We tabulated the use of other support devices, including wheelchair usage, intrathecal baclofen pumps, tracheostomy, percutaneous gastrostomy, hearing aids and ventriculoperitonal shunts. Actograms from all three populations were analyzed identically. Partial-day tail recordings were discarded, resulting in an integer number of full “days” (defined as midnight to midnight). Actograms from all three datasets did not contain any missing values. RAR amplitude, mesor and acrophase were computed through a standard cosinor regression. IV (intradaily variability) and IS (intradaily stability) were calculated on hourly totals from open-source code (github.com/wadpac/GGIR). F (or the *pseudo-F statistic*^23,25^), a measure of RAR robustness, was calculated using an extended sigmoidally transformed cosine model (https://github.com/JessLGraves/RAR). Since recording durations varied across subjects, and since longer recordings alone can enhance F^26^, our reported F scores are normalized by recording duration. “Sleep” bouts (enquoted, as these are *estimates* of sleep) were defined as bouts of zero activity lasting at least four minutes^26^. All graphs, frequency distribution histograms, survival curves and statistical comparisons were generated using Prism GraphPad 10. Three-group comparisons (e.g., Fig. 1B) were conducted using an ordinary one-way ANOVA, with post-hoc comparisons through the Tukey’s multiple comparison test. Smoothed actograms (e.g., Fig. 2A) were generated by calculating an ‘average of averages’ (averaging actograms across days and across subjects *within* each cohort), and smoothing this curve by devising a second-order polynomial *after* averaging across 100 nearest neighbors^26^. For *k*-means clustering (Matlab), we assimilated each subject’s Z-normalized total daily “sleep”, amplitude, acrophase and duration-normalized F scores into a single vector. Clustering was conducted using 2000 replicates to return a solution with the lowest total sum of distances.

**Figure 2.**
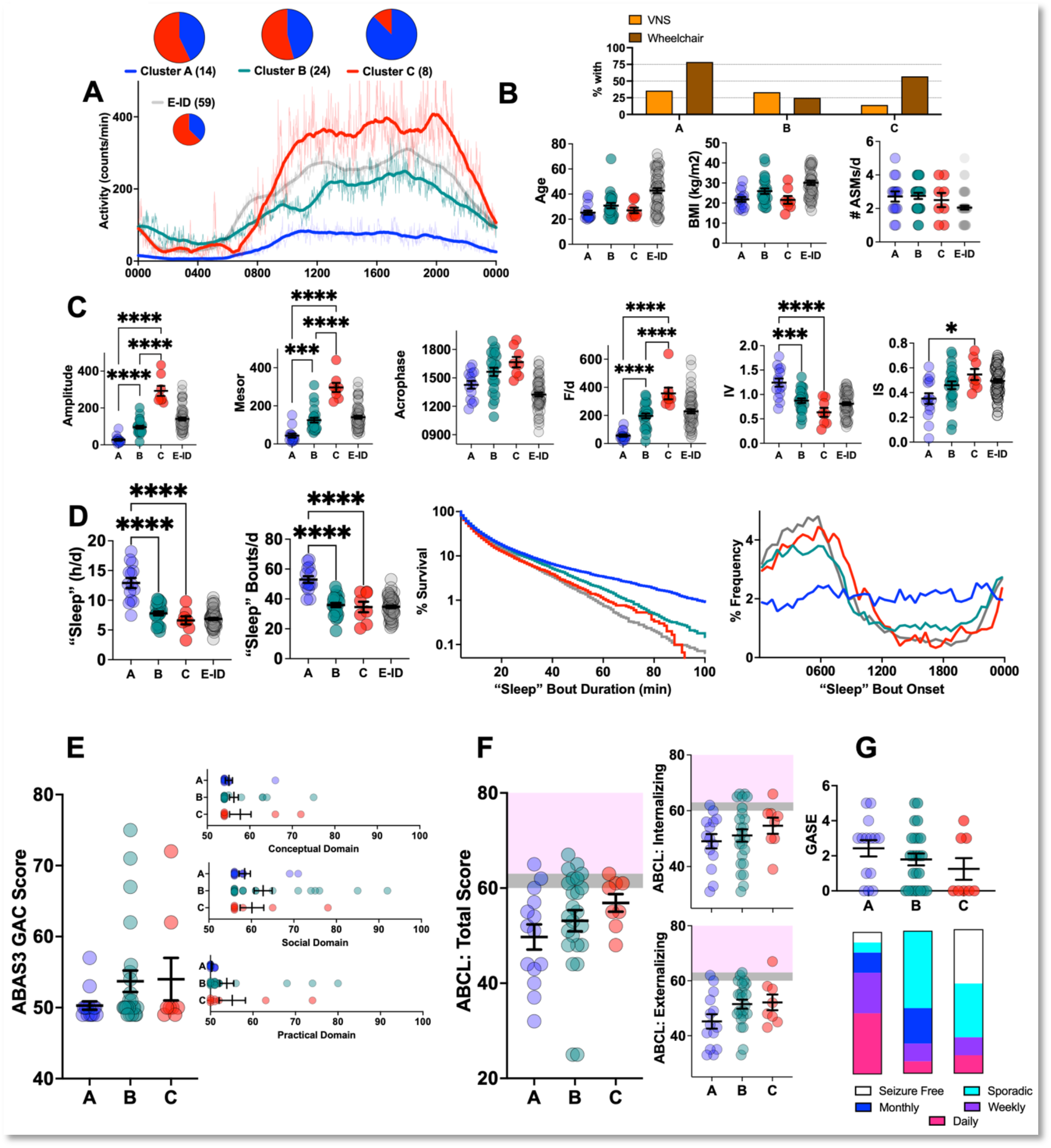
RAR Diversity within E+ID subjects. A: Averaged (thin lines), and averaged *and* smoothed (thick lines) actograms from E-ID subjects divided into three actigraphically defined E+ID clusters defined achieved through *k*-means clustering. Pie charts of sex (blue-male, red-female) shown in inset. B: Proportions of subjects with VNS and wheelchair dependence. Clusters A, B and C were similar in age, BMI and ASM polytherapy. C: RAR parameters from E-ID subjects are shown as a comparison but are not included in ANOVA calculations. Clusters varied significantly by amplitude (F2,43 = 96, p<0.0001), mesor (F2,43 = 50.7, p<0.0001), acrophase (F2,43 = 4.5, p<0.05), F/d (F2,43 = 37.3, p<0.0001), IV (F2,43 = 15.2, p<0.0001) and IS (F2,43 = 4.5, p<0.05). Cluster A subjects displayed the weakest RARs, and cluster C displayed hyper-robust high amplitude RARs, with cluster B forming an intermediate group. D: Cluster A subjects displayed significantly higher total daily “sleep” (F2,43 = 29.2, p<0.0001) and mean daily “sleep bouts” (F2,43 = 23.7, p<0.0001). Distributions of “sleep” bout durations for three clusters diverge beyond bout durations of ∼40 minutes. “Sleep” bouts in cluster A subjects occurred with equal likelihood throughout the day-night cycle. E: ABAS-3 composite and subscores. F: ABCL total scores and domain subscores. Grey shading depicts “borderline” scores. G: Distribution of GASE scores and variations in seizure frequency. Abbreviations: F/d (duration normalized pseudo-F statistic), IS (interdaily stability), IV (intradaily variability). ABAS3 (Adaptive Behavior Assessment System-3), ABCL (Adult Behavior Check List), GASE (Global Assessment of Severity of Epilepsy). *, **, *** and **** correspond to p<0.05, <0.01, 0.001 and 0.001 for post hoc comparisons. Mean + standard error of the mean shown for all.

## Results

We present data from 46 E+ID subjects. An additional 7 subjects were excluded due to technical error (1) and either incomplete surveys and/or device intolerance (6). Study packets were not returned from an additional two subjects. Table 1 provides a summary of demographic data, epilepsy and EEG-related variables, as well as GASE, ABAS3 and ABCL scores for all E+ID subjects ranked by the burden of ASM polytherapy. ABAS-3 scores reflected low to extremely low^42^ levels of adaptive functioning overall, with E+ID subjects displaying a mean GAC of 52.7 (standard deviation, 6.6). Similar scores were reported in a recent trial of thalamic neuromodulation for adults with Lennox Gastaut Syndrome^45^. Our E+ID cohort also displayed objective evidence of high epilepsy severity, with active vagus nerve stimulation (VNS) in 30% of subjects and seizure freedom in less than 10%. ∼71% of E+ID subjects reported seizure onset prior to age 10. E+ID subjects consumed a mean 2.7 ASMs/d, in contrast to E-ID subjects (∼2.1 ASMs/d) where only ∼10% were receiving VNS^26^. ∼47% of E+ID subjects were wheelchair dependent.

**Table 1.**
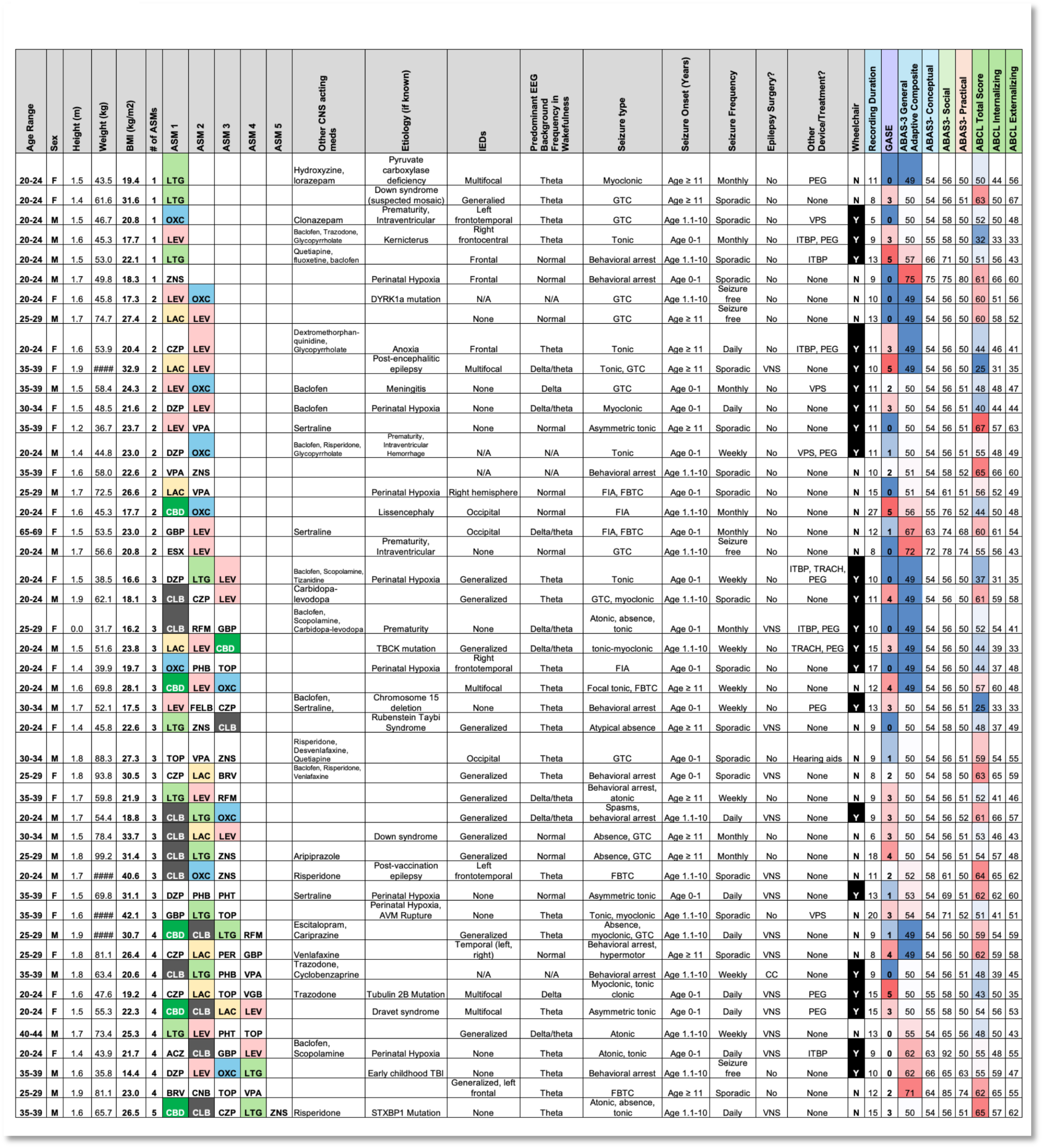

The landscape of ASM prescriptions in both groups is shown in Fig. 1A, which also shows that compared with E-ID and SOL, E+ID subjects had a significantly lower body mass index and provided significantly longer recording durations. Compared with age- and sex-matched SOL subjects, E+ID subjects displayed RARs with deficits in height, robustness and stability. Across most indices, E-ID subjects displayed values intermediate to E+ID and SOL (Fig. 1B, C). E+ID subjects also displayed the highest amounts of total daily “sleep” (Fig. 1D), was achieved primarily through an increase in the number of “sleep” bouts. The distribution of “sleep” bout durations for each group is shown as survival curves (representing the percentage of bouts that *survive* to a particular duration^26,52^). 90% of all sleep bouts in SOL subjects were 21 minutes or shorter, compared with 24 minutes (E-ID) and *28* minutes in E+ID subjects (Fig. 1E). “Sleep” bouts in E+ID subjects were poorly consolidated to a main sleep period, unlike E-ID and SOL subject counterparts (Fig. 1E).

Using four non-collinear actigraphic parameters (see Methods), we *k-*means clustered E+ID subjects into three subgroups with distinct RAR phenotypes. Recognized clusters were similar in age, BMI and ASM polytherapy (Fig. 2A-D). Cluster “A” subjects displayed the weakest RARs, with marked deficits in amplitude and robustness, together with high rates of intradaily variability. This cohort also displayed the highest measures of total daily sleep, accomplished through a greater number of “sleep” bouts that were somewhat uniformly distributed throughout the day. Most cluster “A” subjects were wheelchair dependent (11/14), and examples of actograms and “sleep” measures for cluster “A” subjects are shown in Fig. 3A-C. Cluster “B” subjects had lower rates of wheelchair dependence (6/24) and exhibited RAR parameters that were comparable to E-ID (Fig. 3D-G). Cluster “C” subjects were mostly male and 5/8 required wheelchair support. This cluster of subjects displayed the most vigorous RARs, with high amplitudes, robustness and interdaily stability (Fig. 3H, I), but with measures of “sleep” that were similar to E-ID and cluster “B”. ABAS-3 and ABCL scores were not significantly different among these groups (Fig. 2E,F). While GASE scores were not significantly different between groups, cluster “C” contained the highest proportion of seizure-free patients (Figure 2G). We illustrate representative actograms from all three clusters in Figure 3.

**Figure 3.**
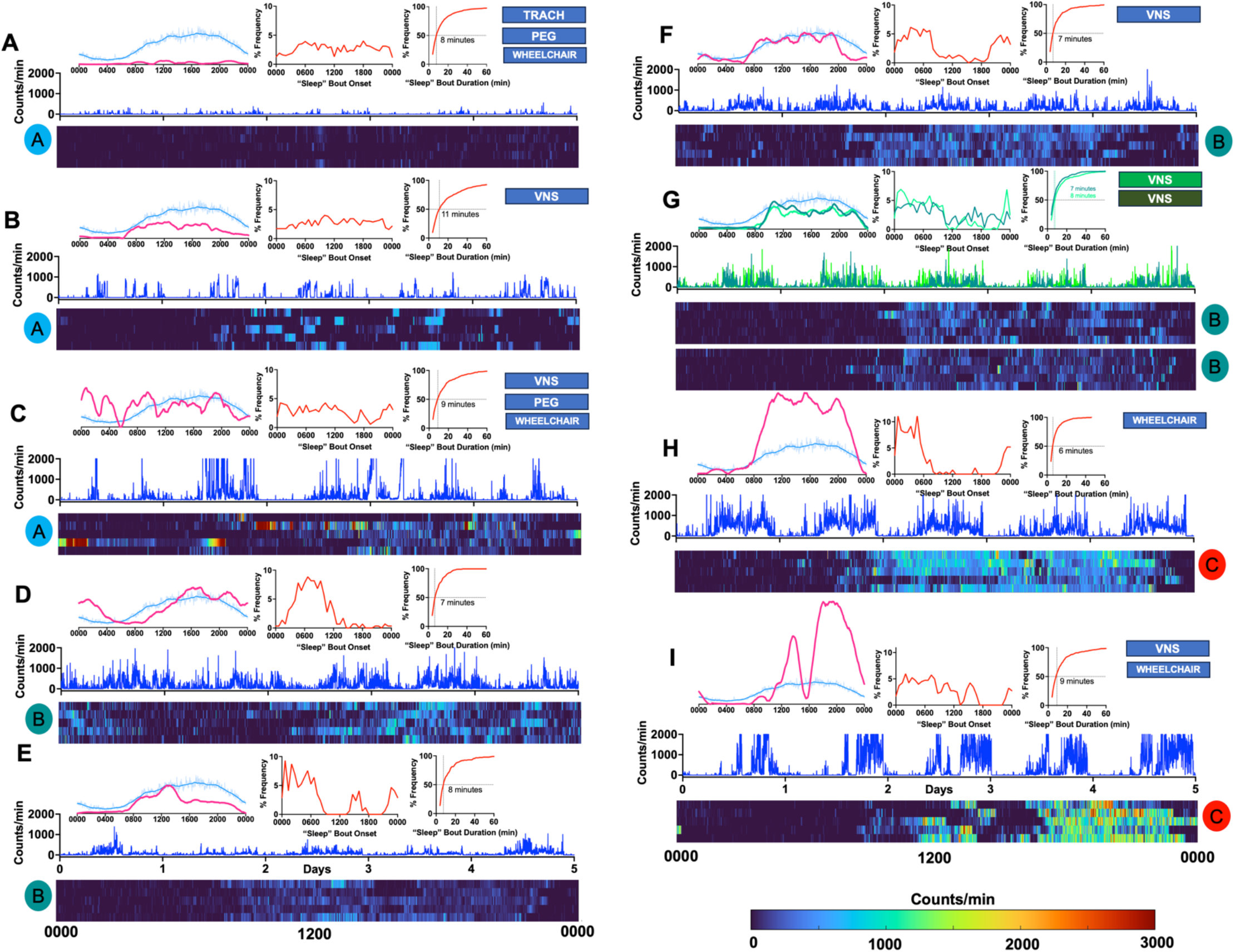
Representative Actograms. Data from each subject is depicted in five sub-panels, with cluster designations shown as colored circles. A smoothed actogram (averaged across all days of recording) for each *individual subject* (pink) is compared against the average smoothed actogram of the entire E+ID cohort (light blue). The distribution of the subject’s “sleep” bout onset and a cumulative frequency distribution histogram of “sleep” bout durations are also shown. The first five days of actigraphy data are depicted as a time series (dark blue, y-axis capped at 2000 counts/min) and also as a heatmap, where every row corresponds to a day (midnight to midnight) and every cell within that row is color coded by relative activity counts (range shown in the bottom right). A: 20-24yo (year old) male patient with a pathogenic *TBCK* mutation (+TRACH, +PEG, wheelchair), with very low levels of sporadic activity detected. B: A 35-39 yo male patient with a pathogenic *STXBP1* mutation (+VNS), where activity levels are higher, but remain sporadic and discontinuous. C: A 20-24yo female patient with Dravet syndrome (+VNS, PEG, wheelchair), illustrating marked day to day variability in RARs. D: A 20-24yo M with known perinatal hypoxia, providing an example of a marked acrophase delay. E: A 30-34yo male patient with Down syndrome. F: A 20-24yo female patient with Rubinstein-Taybi syndrome (+VNS). G: Actograms from two female patients that are 25-29yo, who are biological siblings (both with VNS). H: A 20-24yo male patient (+wheelchair). I: A 20-24yo male patient (+ VNS, wheelchair), illustrating a relatively consistent mid-day nap period. Abbreviations: VNS (vagus nerve stimulation), TRACH (tracheostomy), PEG (percutaneous gastrostomy).

## Discussion

Within any adult epilepsy clinic, patients with comorbid ID represent a medically vulnerable subgroup. From the perspective of seizure control, E+ID persons display higher rates of *both* drug refractoriness *and* intolerable ASM neuropsychiatric side effects^7^. By virtue of their early onset epilepsy, they are subjected to the uncertainties surrounding pediatric to adult transitions of care. They experience the highest rates of CNS polypharmacy, somatic multimorbidity and neurobehavioral comorbidities that frequently necessitate institutionalization. E+ID persons also display the highest rates of premature mortality^13^, including via sudden unexpected death in epilepsy^14,53^. With continued investments in aggressive early medical/surgical seizure prevention strategies, and with the increasing utilization of life-sustaining supportive therapies in childhood (e.g., tracheostomies, gastrostomies), the fraction of E+ID subjects in adult epilepsy clinics is expected to continue to expand. Since deficits in social communication and language frequently accompany moderate to severe ID, seizure burden and neuropsychiatric wellbeing must often be deduced or inferred from subjective caregiver reports. Periodic neurological examinations in ambulatory clinic settings may under- (or over-) estimate the extent of circadian and sleep disruptions, hyperactivity and challenging behaviors, or impairments in consciousness/static encephalopathy seen at home. For these many reasons, E+ID patients and their caregivers are most likely to benefit from technological innovations that non-obtrusively provide objective and holistic correlates of neuropsychiatric wellbeing that are captured in a patient’s native environment.

To this end, we applied wrist actigraphy to describe rest-activity rhythms (RARs) and sleep abnormalities in persons with E+ID. Our results confirm the general feasibility and acceptability of this approach, and we report two main findings. First, compared with E-ID and SOL control cohorts, E+ID subjects overall displayed significant reductions in both RAR height (amplitude/mesor) and stability (low IS, F, and high IV). Second, *within* our E+ID cohort, we recognized marked heterogeneities in RAR morphology. As an initial attempt to classify these RAR morphologies in an unsupervised fashion, we applied *k-*means clustering to identify a cohort with marked hypersomnia and inactivity (A), a cohort with extremely robust and hyperactive RARs (C), and an intermediate cluster (B) with RAR parameters like E-ID subjects. Future studies that incorporate larger sample sizes will be necessary to clarify the practical value of this preliminary clustering approach. Importantly, all three subject clusters had similar ABAS3 and ABCL scores, indicating that our RAR metrics capture a phenotype that is poorly correlated with standardized scores of adaptive function or comorbid psychiatric symptomatology.

We acknowledge several limitations. First, as with any cross-sectional actigraphic evaluation, pinpointing the determinants of such aberrant RAR profiles is challenging^31^. Deficits in RAR amplitude may reflect the extent of an underlying encephalopathy (epileptic or other), the psychomotor retardation associated with ASM polytherapy, and/or frank motor weakness^39^ (quadriparesis). In contrast, comparatively high activity counts may result from motor stereotypies, tics and/or myoclonus. Similarly, individual differences in RAR acrophase may be secondary to caregiver-imposed routines, rather than a measure of intrinsic chronotype (as shown in Fig. 3G, where RARs obtained from two co-habiting sisters were quite similar in timing and amplitude). Thus, while the precise causes of specific RAR abnormalities may be impossible to ascertain, understanding the long-term stability or volatility of such RAR abnormalities may have more practical value. Recent innovations in battery life and wireless data transfer now provide an opportunity to conduct weeks- or months-long actigraphy recordings^54-57^. Incorporating such longitudinal assessments will be essential to define the precise contexts of use for wrist actigraphy in E+ID populations. As an example, continuous long-term measurements of total daily sleep and RAR amplitude may provide an opportunity to individualize ASM dosing to avoid toxicity, and/or assess recovery following health deteriorations (e.g., seizure exacerbations, pneumonia). While the sampling rate of activity counts in actimeters (e.g., Actiwatch-2) may be insufficient to enable accurate seizure detection, wrist actigraphy may be more well suited to depict the magnitude and duration of post-ictal inactivity. Continuous actigraphy recordings may also provide useful feedback when implementing behavioral modifications to intentionally restore aberrant RARs, such as through scheduled activity times^58^. In addition to boosting robustness through routinization, such RAR-adjusting treatments could potentially align a subject’s maximally wakeful periods with behavioral, physical and occupational therapies designed to improve adaptive function.

Second, our relatively small sample was derived from a tertiary care epilepsy referral center where rates of medically intractable seizures tend to be higher. Our E+ID subjects were prescribed an average of 2.7 daily ASMs, and over ∼30% of E+ID subjects were receiving VNS therapy. While all caregivers that we approached were enthusiastic about the goals of the study, several declined to consent out of concerns for device intolerability (“he/she just *can’t/won’t* wear a watch”, “he/she will try to eat or bite the watch”). We also did not seek consent from official state-appointed guardians of orphan adult E+ID patients who were typically accompanied by a representative from their group home. These elements may have resulted in a selection bias, effectively oversampling subjects with more virulent epilepsies, while under sampling those with challenging behaviors and absent familial caregivers.

Third, we adopted a fairly simple algorithm^26^ to estimate bouts of “sleep” without incorporating any diary data pertaining to bed times and wake times. This may have incorrectly designated certain daytime epochs of quiet wakefulness as “sleep” in subjects with profound motor weakness. Discerning state/stages of sleep using wrist-worn data alone has been achieved by combining actigraphy data with simultaneous electrocardiography^59-61^ (sampling heart rate and heart rate variability). However, these machine-learning based classifier models may require further adjustments in E+ID populations, as they are frequently trained on neurotypical subjects (with or without subjective sleep complaints).

In conclusion, our study provides a cross-sectional RAR characterization of a dedicated sample of adults with epilepsy and intellectual disability. By comparing these metrics against a sample of adults with epilepsy without intellectual disability (E-ID) and a historically collected set of age- and sex-matched control subjects, we spotlight the profound deficits in RAR height and robustness observed in E+ID. By incorporating RAR endpoints as monitoring biomarkers^62^ into routine patient care and drug/device treatment trials, we may sensitively detect small but clinically meaningful improvements in activity levels and sleep/sleep timing^15^, when traditional survey-based clinical outcome assessments (e.g., ABAS-3) maybe insufficiently sensitive. Recognizing the full potential of wrist actigraphy in E+ID populations will require (i) prospective longitudinal evaluations with contemporaneously annotated health/symptom data, and (ii) improvements in device form factors that allow for seamless data collection in individuals with sensory processing symptoms that preclude continuous watch-wearing.

## Data Availability

All raw data from E+ID and E-ID subjects in the current study are available upon reasonable request. Data from subjects in the SOL (Study of Latinos) study can be downloaded with permission from the National Sleep Research Resource (www.sleepdata.org).

## Author contributions

All authors satisfy the ICMJE guidance by substantially contributing to the design, analysis and interpretation of the work, drafting of the manuscript, final approval of the manuscript and all agree to be accountable for all aspects of the work in ensuring that questions related to the accuracy or integrity of any part of the work is appropriately investigated and resolved.

